# Validation and Refinement of Operator Syndrome in Active-Duty Special Operations Forces

**DOI:** 10.1101/2025.11.06.25339692

**Authors:** Shane W. Adams, B. Christopher Frueh, Jerome Sabangan, Carey Pawlowski, Robert Oh, Cameron Pugach, Odette Harris

**Affiliations:** Polytrauma System of Care, Veterans Affairs Palo Alto Health Care System, Palo Alto, CA, USA; Department of Neurosurgery, Stanford University School of Medicine, Palo Alto, CA, USA; Department of Psychology, University of Hawaii – Hilo, Hilo, HI, USA; Department of Medicine, Stanford University School of Medicine, Palo Alto, CA, USA; Behavioral Science Division, National Center for PTSD at VA Boston Healthcare System, Boston, Massachusetts, USA; Department of Psychiatry, Boston University Chobanian & Avedisian School of Medicine, Boston, Massachusetts, USA

**Keywords:** Special Operations, Operator Syndrome, Integrative, Whole Health, Systemic, Traumatic Brain Injury, Stress, Polytrauma

## Abstract

**Background:** Operator Syndrome (OS) describes the unique constellation of symptoms observed in special operations forces (SOF) following a) chronic and repetitive exposure to high-tempo tasks that are physically demanding and involve extreme threats to physical safety, b) are compounded by potentially traumatic events, and/or c) traumatic brain injury (TBI). This study sought to empirically test the validity of OS and refine OS to reflect the latest evidence.

**Methods:** Active-duty, treatment seeking SOF with significant histories of TBI who participated in VA Palo Alto’s Intensive Evaluation and Treatment Program (*n* = 202) completed a battery of multimodal and multidimensional assessments as part of their clinical care. OS was evaluated according to the Middle-Out Approach, using both variable-centered (exploratory factor analysis [EFA]) and person-centered (latent class analysis [LCA]) methods with a closer examination using LCA of unique mental health presentations.

**Results:** A one-factor and one-class structure across 9 domains of OS was indicated by EFA and LCA, respectively. Meeting ≥ 5 domains (Headaches [.679] Mental Health [.634], Pain [.560], Sensory [.524], and Sleep [.441] Disruptions) accounted for 68.9% of the variance in OS outcomes, suggesting a suitable criterion for detecting OS, which was experienced by 180 (89.1%) participants. Unique mental health presentations of Dysphoric Arousal (47.5%) and Hyperarousal (28.7%) were more common than traditionally recognized presentations of anxiety, depression, and posttraumatic stress disorder (23.8%).

**Conclusions:** This study provides the first empirical validation of OS as a unitary construct, as well as evidence of the core features of OS and evidence that OS can provide unique utility beyond current psychiatric diagnoses. Criterion A-C for detecting OS are recommended. Further precision in assessing the systemic influences of OS can help triage effective integrative treatments for OS in SOF personnel.

---

Special operations forces (SOF) are military personnel who pass a highly restrictive selection process and receive advanced training to conduct high-risk and often clandestine unconventional warfare (1,2). United States’ SOF personnel have been heavily relied upon in post-9/11 warfare, the global war on terrorism (GWOT), and now, post-GWOT to accomplish the most daring and difficult military tasks across a range of missions (e.g., direct action, hostage rescue, reconnaissance, counterterrorism, counterinsurgency, civil affairs) (2). The men and women who carry out these missions are known as “operators,” a term reflecting the precision and professionalism with which these tasks are meant to be completed.

In 2004, U.S. Special Operations Command (USSOCOM) was given the broader objective of leading combat-related efforts in GWOT (3). To meet this objective, U.S. SOF personnel grew from approximately 38,000 in 2001 to 70,000 in 2025 (3). Despite comprising less than 3% of the conventional military, SOF are often relied on to carry out the most critical missions that shape our geopolitical landscape (2). Increasing reliance and responsibility demands more frequent and chronic high-tempo training and deployment operations that can ultimately produce “a cumulative effect on the force,” according to a USSOCOM comprehensive review (4). As an unintended result, many SOF can incur a greater burden of health-related consequences compared to conventional military forces (5–7). Although governing and healthcare agencies have increasingly recognized the cumulative health-related costs of chronic high-tempo SOF operations, more progress is needed to meet the emerging healthcare needs of SOF personnel (5–7). Most notably, more research is needed to identify and evaluate the breadth of complex injuries and related symptoms, their correlates and collateral systemic effects, as well as unique subpopulations within SOF that may require more individualized care.

## Operator Syndrome

The unique constellation of postconcussive, behavioral, biological, and physiological sequelae experienced by SOF have come to be collectively termed, Operator Syndrome (OS) (5). In addition to traumatic brain injury (TBI), the original OS conceptual model included symptom domains of chronic pain/headaches, cognition, endocrine, mental health, perceptual system, sleep, substances, marital/relational, toxin exposures, military transition, and existential concerns (5). OS can result from chronic and repetitive exposure to high-tempo tasks that are physically demanding, involve extreme threats to physical safety, and are compounded by potentially traumatic events and/or TBI. These exposures are not necessarily specific to SOF operators but are more common to SOF operators because of the extreme demands of their training, deployment, and accompanied lifestyle. Therefore, OS provides a useful framework to guide evaluation and treatment practices for the cumulative effects of multiple physical (i.e., TBI, musculoskeletal) and psychological (i.e., trauma, chronic stress/allostatic load) injuries (5).

## Polytrauma System of Care

The framework of OS does well to capture the breadth of injuries and related symptoms observed in many service members and veterans receiving care from Veterans Health Administration (VHA) Polytrauma System of Care (PSC) (8). PSC was created in 2004 to address complex patterns of multiple injuries during the course of GWOT, which primarily include TBI, other bodily injuries (e.g., musculoskeletal, amputation), and related physical, sensory, cognitive, or psychological symptoms. These injuries and sequelae parallel OS and require an extraordinary level of integrated and coordinated healthcare services, central to the PSC’s clinical model. Moreover, within PSC, the Intensive Evaluation and Treatment Program (IETP) was created specifically to serve active-duty and veteran SOF personnel following these injuries, making IETP and PSC optimal settings to evaluate and treat OS.

As with all PSC patients, precision is needed to understand the nuances of complex multiple injuries and to identify subpopulations that may require further personalized treatments (9,10). This aim is particularly relevant to SOF. Rather than trying to assess and treat relevant complex health issues from a conventional model of care or a one-size-fits-all model that may fail to fully and accurately describe an individual’s unique presenting issues, their extent, and comprehensive avenues for treatment, a precision rehabilitation approach is needed to integrate a broader range of health information (e.g., behavioral, biological, *and* physiological) to help researchers and clinicians identify and address the unique constellations of complex injuries and sequelae relevant to OS. While PSC and IETP provide the optimal interdisciplinary setting and resources to assess and treat OS within a comprehensive and integrative care model, the Middle-Out Approach (MOA) provides complementary methods to evaluate and bring precision to the study of OS (10).

## Middle-Out Approach

The primary aim of MOA is to empirically evaluate, bring precision to, and represent unique and complex health experiences that may not be fully captured by current diagnoses, assessments, and treatments (10). Therefore, MOA is best suited to facilitate the mission of PSC/IETP, evaluate OS, and bring precision to both to better meet the emerging needs of SOF and other service members and veterans. To do this, MOA uses both variable- and person-centered statistical analyses across a range of transdiagnostic, multidimensional, and multimodal variables to address the methodological complexities of identifying increasingly precise clinical phenotypes that may be unique to specific individuals or communities, like SOF, as well as unique subpopulations within those communities (see Adams et al. for more details) (10).

## Current Study

Frueh et al.’s (5) conceptual framework was an essential first step in deriving a working definition of OS and gathering feedback from clinicians, researchers, and the SOF community. However, there are limitations that must be addressed to continue the development of OS. First, there is no *quantitative* empirical evidence to support the validity of OS. Formal clinical syndromes and diagnoses must not only have clinical utility and support from the community but also sufficient empirical evidence and replication across studies with strong methodological rigor. Without validation, the development of OS and any adoption to policy and implementation practices is stymied. Second, critics may contend that OS does not add additional clinical utility beyond existing diagnoses like posttraumatic stress disorder (PTSD) or already recognized TBI-related sequelae. Formal clinical syndromes and diagnoses must have strong rationale, supporting evidence, clarity and scope, and expected impact on clinical practice and public health beyond current practices (11). Part of this work includes identifying the core features of OS to limit the scope and potential overlap with existing diagnoses.

To address current limitations, this study 1) empirically tested the validity of OS using MOA in a sample of treatment-seeking active-duty SOF personnel; 2) refined OS to reflect its core features and developed clear criteria for the assessment of OS; and 3) identified precise symptom presentations of OS mental health disruption that can help parse OS apart from existing psychiatric diagnoses like anxiety, depression, and PTSD. Lastly, we evaluated preliminary data regarding additional clinically-relevant OS domains (e.g., moral injury, heavy metals/toxin exposure) for future study, and provide recommendations for research, clinical practice, and policy.

## Methods

### Participants and Procedures

Participants included patients who participated in VA Palo Alto’s IETP between 2021-2025. IETP is a three-week residential rehabilitation program housed within the Polytrauma Transitional Rehabilitation Program at the five national VHA PSC sites (i.e., Minneapolis, Palo Alto, Richmond, San Antonio, Tampa). To participate in IETP, patients must be active-duty or veteran SOF and have a significant history of TBI. Participants completed an intensive interdisciplinary treatment composed of medical, psychology, neuropsychology, social work, nursing, occupational therapy, recreational therapy, physical therapy, speech pathology, nutrition, and other consults as needed (e.g., pain management, endocrinology, audiology). All measures in this study were collected during the course of clinical care at VA Palo Alto. Fasting venous blood serum samples were obtained by venipuncture between 0700 and 0800 for each participant, and samples were immediately analyzed by the VA clinical lab or sent to Quest Diagnostics.

To date, 246 patients have completed IETP Palo Alto. The first 44 participants of the program were excluded because their participation preceded program implementation of the current measures. This resulted in a final sample of 202 participants. Examination of participants excluded (*n* = 44) and included (*n* = 202) indicated no significant differences in anxious, depressive, posttraumatic stress, sleep, nor neurobehavioral symptoms, suggesting minimal selection bias in this final sample. Procedures of this study were approved by the VHA-affiliated Stanford University Institutional Review Board (Protocol #17671).

### Measures

All measures were collected at baseline by the relevant clinician (e.g., psychologist— depressive symptoms, physical therapist—sensory disruption). See Table 1 for a list of measures and Table 2 for cutoff scores used as domain criteria, which were based on referenced norms for clinical significance. Below we provide a brief overview of all measures by OS domain. Please see references for more details.

**Table 1.** Operator Syndrome Domains and Measures Used in IETP.

| **Domain and Measure** | **Abbreviation** |
| --- | --- |
| ***Cardiometabolic Disruption*** |  |
| Body Mass Index | BMI |
| Blood Pressure (systolic/diastolic) | BP |
| Cholesterol Total | Chol |
| Glucose Blood | Gluc |
| High-Density Lipoprotein Cholesterol | HDL |
| Low-Density Lipoprotein Cholesterol | LDL |
| Triglycerides | Tri |
| ***Chronic Localized and Non-Specific Pain*** |  |
| Non-Specific Posttraumatic Somatic Distress | NSPSD |
| Pain Catastrophizing Scale | PCS |
| Patient-Reported Outcomes Measurement Information System – Pain Interference | PROMIS |
| ***Cognitive Disruption*** |  |
| Cognitive-Linguistic Quick Test | CLQT |
| Design Generation | DG |
| Neuro-QoL Cognitive Function | Neuro-QoL |
| Test of Memory and Learning | TOMAL |
| Digits Forward | DF |
| Paired Recall | PR |
| Word Selective Reminding | WSR |
| Word Selective Reminding Delayed Recall | WSRD |
| ***Endocrine Disruption*<sup>a</sup>** |  |
| Total Follicular Stimulating Hormone | FSH |
| Total Luteinizing Hormone | LH |
| Total Testosterone | T |
| Total Thyroid Stimulating Hormone | TSH |
| ***Gastrointestinal Disruption*** |  |
| Clinical Interview (e.g., chronic constipation, GERD, IBS, abdominal cramping) | - |
| ***Headaches*** |  |
| Headache Impact Test-6 | HIT-6 |
| Migraine Disability Assessment Scale | MIDAS |
| ***Heavy Metals/Toxins*** |  |
| Total Blood Serum |  |
| Arsenic | As |
| Lead | Pb |
| Manganese | Mn |
| Mercury | Hg |
| Total Random Urine |  |
| Arsenic | As |
| ***Mental Health Disruption*** |  |
| General Anxiety Disorder-7 | GAD-7 |
| Patient Health Questionnaire-9 | PHQ-9 |
| PTSD Checklist for DSM-5 | PCL-5 |
| ***Moral Injury, Existential Disruption, and Military Transition*** |  |
| Moral Injury and Distress Scale | MIDS |
| ***Sensory Disruption*** |  |
| Benign Paroxysmal Positional Vertigo | BPPV |
| Dizziness Handicap Inventory | DHI |
| Sensory Organization Test (Somatosensory, Visual, Vestibular) | SOT |
| Speech, Spatial, and Qualities of Hearing Scale | SSQ-12 |
| ***Sleep Disruption*** |  |
| Insomnia Severity Index | ISI |

**Table 2.**
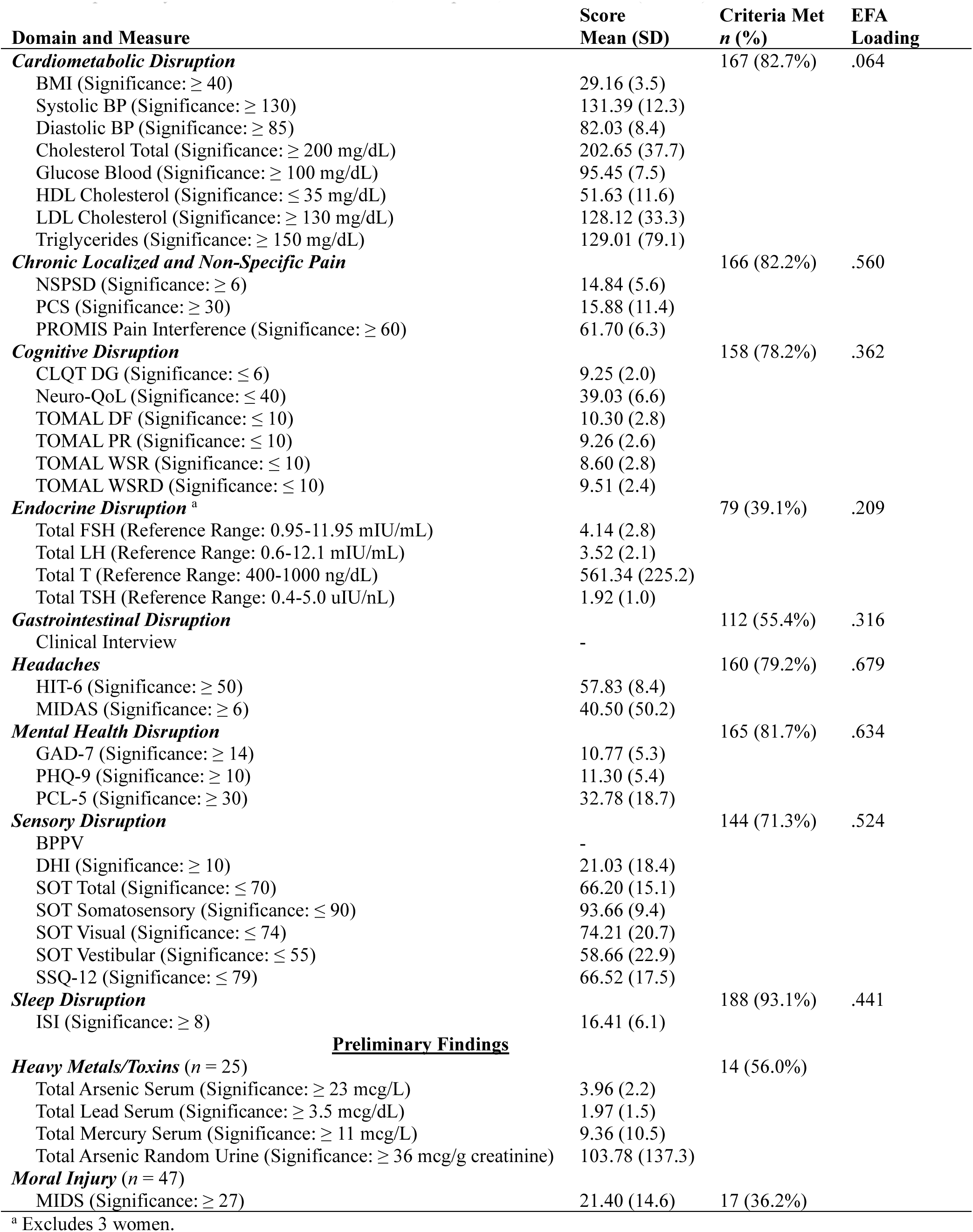
Operator Syndrome Domains Examined, Descriptives, and EFA results (*n* = 202)

| Domain and Measure | Score<br>Mean (SD) | Criteria Met<br>$n$ (%) | EFA<br>Loading |
| --- | --- | --- | --- |
| **Cardiometabolic Disruption** |  | 167 (82.7%) | .064 |
| BMI (Significance: $\geq 40$) | 29.16 (3.5) | | |
| Systolic BP (Significance: $\geq 130$) | 131.39 (12.3) | | |
| Diastolic BP (Significance: $\geq 85$) | 82.03 (8.4) | | |
| Cholesterol Total (Significance: $\geq 200$ mg/dL) | 202.65 (37.7) | | |
| Glucose Blood (Significance: $\geq 100$ mg/dL) | 95.45 (7.5) | | |
| HDL Cholesterol (Significance: $\leq 35$ mg/dL) | 51.63 (11.6) | | |
| LDL Cholesterol (Significance: $\geq 130$ mg/dL) | 128.12 (33.3) | | |
| Triglycerides (Significance: $\geq 150$ mg/dL) | 129.01 (79.1) | | |
| **Chronic Localized and Non-Specific Pain** |  | 166 (82.2%) | .560 |
| NSPSD (Significance: $\geq 6$) | 14.84 (5.6) | | |
| PCS (Significance: $\geq 30$) | 15.88 (11.4) | | |
| PROMIS Pain Interference (Significance: $\geq 60$) | 61.70 (6.3) | | |
| **Cognitive Disruption** |  | 158 (78.2%) | .362 |
| CLQT DG (Significance: $\leq 6$) | 9.25 (2.0) | | |
| Neuro-QoL (Significance: $\leq 40$) | 39.03 (6.6) | | |
| TOMAL DF (Significance: $\leq 10$) | 10.30 (2.8) | | |
| TOMAL PR (Significance: $\leq 10$) | 9.26 (2.6) | | |
| TOMAL WSR (Significance: $\leq 10$) | 8.60 (2.8) | | |
| TOMAL WSRD (Significance: $\leq 10$) | 9.51 (2.4) | | |
| **Endocrine Disruption<sup>a</sup>** |  | 79 (39.1%) | .209 |
| Total FSH (Reference Range: 0.95-11.95 mIU/mL) | 4.14 (2.8) |  |  |
| Total LH (Reference Range: 0.6-12.1 mIU/mL) | 3.52 (2.1) |  |  |
| Total T (Reference Range: 400-1000 ng/dL) | 561.34 (225.2) |  |  |
| Total TSH (Reference Range: 0.4-5.0 uIU/nL) | 1.92 (1.0) |  |  |
| **Gastrointestinal Disruption** |  | 112 (55.4%) | .316 |
| Clinical Interview | - |  |  |
| **Headaches** |  | 160 (79.2%) | .679 |
| HIT-6 (Significance: $\geq 50$) | 57.83 (8.4) | | |
| MIDAS (Significance: $\geq 6$) | 40.50 (50.2) | | |
| **Mental Health Disruption** |  | 165 (81.7%) | .634 |
| GAD-7 (Significance: $\geq 14$) | 10.77 (5.3) | | |
| PHQ-9 (Significance: $\geq 10$) | 11.30 (5.4) | | |
| PCL-5 (Significance: $\geq 30$) | 32.78 (18.7) | | |
| **Sensory Disruption** |  | 144 (71.3%) | .524 |
| BPPV | - |  |  |
| DHI (Significance: $\geq 10$) | 21.03 (18.4) | | |
| SOT Total (Significance: $\leq 70$) | 66.20 (15.1) | | |
| SOT Somatosensory (Significance: $\leq 90$) | 93.66 (9.4) | | |
| SOT Visual (Significance: $\leq 74$) | 74.21 (20.7) | | |
| SOT Vestibular (Significance: $\leq 55$) | 58.66 (22.9) | | |
| SSQ-12 (Significance: $\leq 79$) | 66.52 (17.5) | | |
| **Sleep Disruption** |  | 188 (93.1%) | .441 |
| ISI (Significance: $\geq 8$) | 16.41 (6.1) | | |
| **Preliminary Findings** |  |  |  |
| **Heavy Metals/Toxins ($n = 25$)** |  | 14 (56.0%) |  |
| Total Arsenic Serum (Significance: $\geq 23$ mcg/L) | 3.96 (2.2) | | |
| Total Lead Serum (Significance: $\geq 3.5$ mcg/dL) | 1.97 (1.5) | | |
| Total Mercury Serum (Significance: $\geq 11$ mcg/L) | 9.36 (10.5) | | |
| Total Arsenic Random Urine (Significance: $\geq 36$ mcg/g creatinine) | 103.78 (137.3) | | |
| **Moral Injury ($n = 47$)** |  |  |  |
| MIDS (Significance: $\geq 27$) | 21.40 (14.6) | 17 (36.2%) | |

<sup>a</sup> Excludes 3 women.

#### Cardiometabolic Disruption

Cardiometabolic disruption reflects a series of systemic biological changes, often as a result of TBI or chronic stress (12,13), that are related to metabolic syndrome, which includes central obesity, elevated blood pressure, lipids, and glucose (14). In this study, elevated body mass index (BMI), systolic and diastolic blood pressure, total blood serum cholesterol, blood serum glucose, blood serum high-density lipoprotein (HDL) and low-density lipoprotein (LDL) cholesterol, and blood serum triglyceride levels were used to indicate broader issues of Cardiometabolic Disruption.

#### Chronic Localized and Non-Specific Pain

The Non-Specific Posttraumatic Somatic Distress (NSPSD) scale, Pain Catastrophizing Scale (PCS), and Patient-Reported Outcomes Measurement Information System (PROMIS) – Pain Interference scale were used to assess pain. The NSPSD scale comprises a list of non-specific pain symptoms that have been used to help predict posttraumatic stress symptoms with a reasonable degree of accuracy (15). The PCS measures how one thinks about their chronic pain (16). The PROMIS – Pain Interference scale measures how much one’s pain interferes with daily activities (17).

#### Cognitive Disruption

The Neuro-QoL Cognitive Function was used to evaluate perceived or subjective cognitive performance issues (18). Objective cognitive performance was evaluated using the Design Generation subtest of the Cognitive-Linguistic Quick Test (CLQT) to assess working memory, problem solving, and executive function (19). Digits Forward, Paired Recall, Word Selective Reminding, and Word Selective Reminding Delayed Recall subtests of the Test of Memory and Learning (TOMAL) were also used to assess memory, attention, and learning (20).

#### Endocrine Disruption

Total blood serum follicular stimulating hormone, luteinizing hormone, testosterone, and thyroid stimulating hormone were used to assess Endocrine Disruption in the current study (21). Considering the potential for higher-than-average baseline testosterone levels in this unique SOF population, a total testosterone threshold ≤ 400 ng/dL was used to detect clinical significance (i.e., testosterone grey zone) (22). Examination of endocrine disruption excluded women due to sex variations.

#### Gastrointestinal Disruption

Gastrointestinal Disruption (e.g., chronic constipation or diarrhea, chronic abdominal cramping, changes in stool frequency/consistency, chronic gastroesophageal reflux) was assessed using clinical interview with participants during their intake process. Disruptions were dichotomously recorded as positive or negative.

#### Headaches

The Headache Impact Test (HIT-6) (23) and Migraine Disability Assessment Scale (MIDAS) (24) were both used to assess the prevalence and functional impact of headaches and migraines.

#### Heavy Metals/Toxins

Regular testing of total blood serum and random urine for heavy metals, including arsenic, lead, and mercury was initiated later in the development of IETP (25). Although this is ongoing for future study and clinical care, currently, a small subset of participants has this data available (*n* = 25). Blood serum and random urine were used to increase sensitivity of arsenic assessment (25).

#### Mental Health Disruption

Anxious, depressive, and posttraumatic stress symptoms were assessed using the General Anxiety Disorder (GAD-7) (26), Patient Health Questionnaire (PHQ-9) (27), and the PTSD Checklist (PCL-5) (28), respectively.

#### Moral Injury and Existential Disruption

Moral injury and existential disruption were assessed using the Moral Injury and Distress Scale (MIDS) (29). The MIDS queries cognitive, emotional, behavioral, social, and religious or spiritual reactions to a range of events that may have conflicted with the participants’ moral beliefs and/or resulted in a jaded change in negative thoughts and existential beliefs about oneself, others, and/or the world in general.

#### Sensory Disruption

The Speech, Spatial, and Qualities of Hearing (SSQ-12) scale was used to assess ability to hear and listen in different scenarios (30). Benign paroxysmal positional vertigo (BPPV), or a sudden onset of dizziness or lightheadedness, was assessed using clinical interview and dichotomously recorded as positive or negative (31). The Dizziness Handicap Inventory (DHI) was used to assess self-reported effects of dizziness on daily life (32). Bertec’s Computerized Dynamic Posturography (CDP) was used to perform the Sensory Organization Test (SOT) to assess somatosensory, visual, and vestibular function (33). Bertec’s CDP uses immersive virtual environments with dual-balance force plate technology to supplement assessment and targeted physical therapy interventions for participants experiencing dizziness, balance problems, and/or motion sensitivity.

#### Sleep Disruption

The Insomnia Severity Index (ISI) was used to assess Sleep Disruption (34).

### Statistical Analyses

Data distributions were examined in SPSS Version 30. Assessment scores were dichotomized depending on the referenced cutoff score to indicate clinical significance. Participants met domain criteria if ≥ 1 measure from each domain surpassed the relevant cutoff score. OS construct validity was evaluated according to MOA (10), including 1) examination of descriptives and frequencies, 2) exploratory factor analysis (EFA), and 3) latent class analysis (LCA).

EFA is a variable-centered statistical technique used to determine if a group of variables reflects one or more factors across a large sample. LCA is a person-centered statistical technique used to determine if there are unique subsets of participants who present with unique profiles within a larger sample. Both EFA and LCA provide complementary information regarding variable cohesion within the larger OS framework (i.e., do these domains reflect a unitary OS construct?) and variations within the broader framework of OS (i.e., do these domains affect different SOF personnel differently?), respectively.

Lastly, to help identify precise profiles of mental health symptoms that may be associated with OS, LCA was used to examine anxiety (GAD-7), depressive (PHQ-9), and posttraumatic stress (PCL-5) symptoms concurrently using full information maximum likelihood estimation in Mplus 8.11 (10). Lower fit indices (Bayesian information criterion [BIC], sample-size adjusted BIC [ssBIC], Akaike information criterion [AIC]), higher entropy values, and statistically significant (*p*<.05) Lo–Mendell–Rubin adjusted likelihood ratio test (LMR-LRT) and bootstrapped likelihood ratio test (BLRT) indicated better model fit (35). A combination of these indices, sufficient class size, theoretical coherence, parsimony, and clinical utility were used to select an optimal model (35,36). To address issues of conditional independence between anxiety, depressive, and posttraumatic stress, the local independence assumption was relaxed by estimating parameters considering the residual associations between indicators and a higher number of random starts (37).

## Results

Participants (*n*=202) were 98.5% men (3 women) aged between 29-56 years (*M*=40.57) who included enlisted personnel (*n*=149, 73.8%), warrant officers (*n*=16, 7.9%), and officers (*n*=37, 18.3%). Participants included personnel from the Navy Sea, Air, and Land (SEAL) Teams (*n*=97, 48.0%), Navy Special Warfare Combat Crewman (SWCC; *n*=22, 10.9%), Army Special Forces (*n*= 74, 36.6%), as well as other units (*n*=9, 4.5%). Military ranks ranged from E-2 to O-8. Participants were 84.6% White (*n*=171), 5.9% (*n*=12) Hispanic, 5.0% (*n*=10) Asian, 2.0% (*n*=4) Black, 1.5% (*n*=3) Native American, and 1.0% (*n*=2) who identified as Other. All participants had a significant history of ≥ mild TBI.

### OS Structure

Means and standard deviations for every measure and prevalence of participants who met criteria for each domain are reported in Table 2. Sleep (93.1%), Cardiometabolic (82.7%), Pain (82.2%), Mental Health (81.7%), Headaches (79.2%), and Cognitive (78.2%) Disruptions were most prevalent.

#### Variable-Centered Approach to OS

EFA was performed on the nine OS domains using direct oblimin rotation. The Kaiser-Meyer-Olkin (KMO) indicated an adequate sample size for analysis (KMO=.620). Bartlett’s test of sphericity (χ^2^=97.08, *p* < .001) suggested sufficient correlation between domains (38). Parallel analysis indicated that only factors with an eigenvalue > 1.30 should be retained. Using this criterion, a one-factor structure of OS was retained (Table 2). Cardiometabolic disruption (.064) accounted for a smaller proportion of variance in the OS factor. The strongest factor loadings were observed for Headaches (.679), Mental Health Disruption (.634), Pain (.560), Sensory Disruption (.524), and Sleep Disruption (.441), suggesting these may be the most central components of OS following chronic stress, trauma, and TBI.

Within the OS factor, five domains accounted for 68.9% of the total variance in OS, with four domains accounting for 58.9% and six domains accounting for 77.8% of the total variance in OS. Using the five-domain criteria for OS, 180 (89.1%) participants met criteria for OS. Point biserial correlations were used to indicate discriminant validity with mental health disorders. OS was significantly correlated with anxiety symptoms (GAD-7, *r_pb_*=.368), depressive symptoms (PHQ-9, *r_pb_*=.435), and posttraumatic stress symptoms (PCL-5, *r_pb_*=.243) as expected given that Mental Health Disruptions are included in OS. However, correlations were only of small-to-medium effect size. Collinearity (*r*≥.70) was not present.

#### Person-Centered Approach to OS

One-to-four class LCA models were examined to identify the optimal solution. Although LRTs were significant for a two-class model, there was decrement in fit indices and poor entropy, suggesting a low probability of sufficient distinction between one and two classes. Accordingly, a one-class model was selected as the optimal solution (Supplemental Table 1). The one-class solution reflected elevated probabilities of all domains with the exception of Endocrine Disruption (Probability=.203; Figure 1).

**Figure 1.**
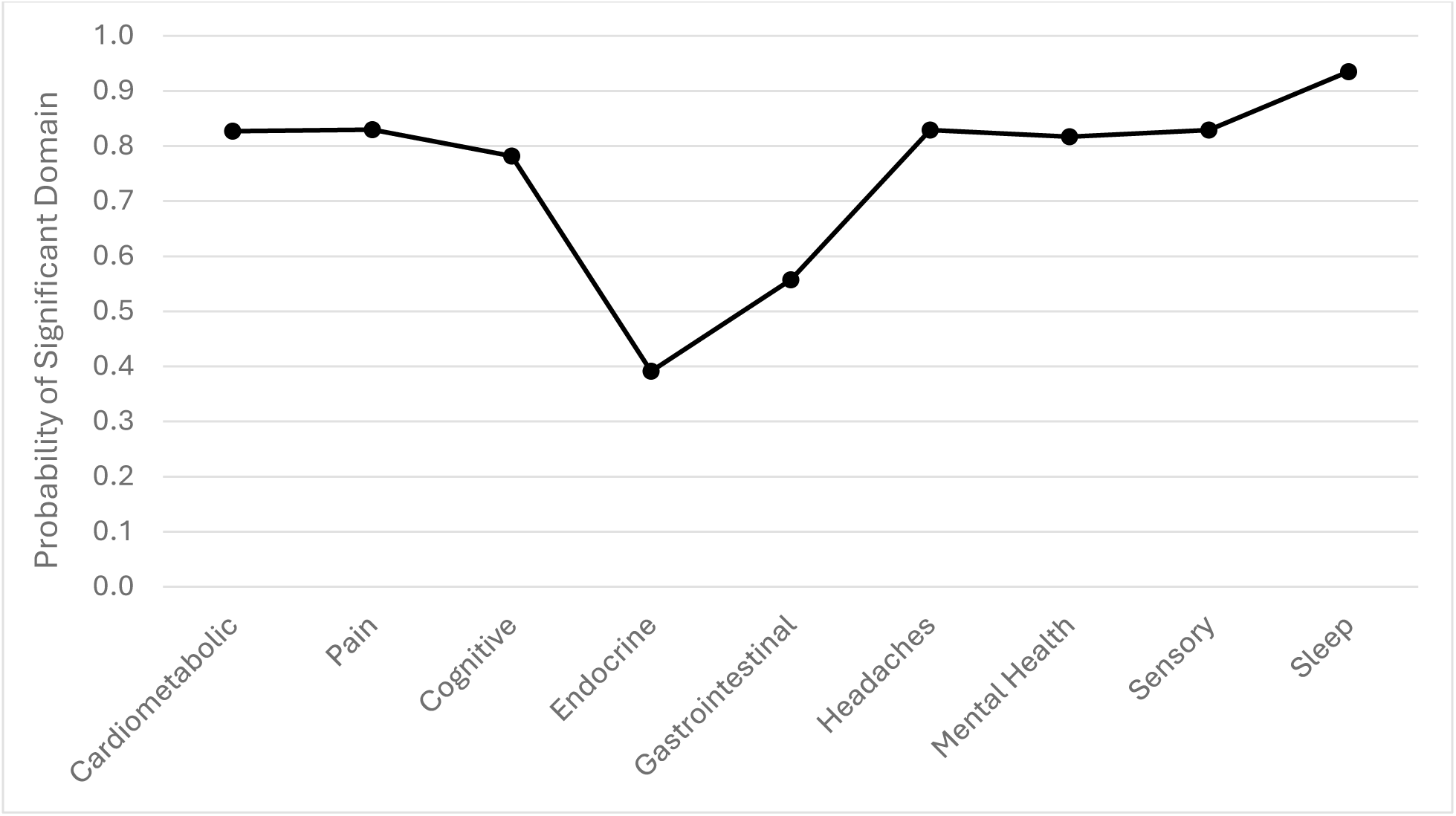
LCA results of one-class OS solution.

### Closer Examination of Mental Health

LCA was conducted on the symptoms of Mental Health Disruption to help curb critiques that contend OS is merely reflective of existing psychiatric diagnoses like PTSD and to identify precise presentations that may provide specific targets for increasingly effective and engaging interventions. Depressive, anxious, and posttraumatic stress symptoms were examined concurrently (10). Two items from the PHQ-9 (concentration, sleep) were excluded from the LCA due to overlap with the PCL-5, resulting in a 34-item LCA.

Using the same selection criteria as above, a three-class optimal solution was selected (Figure 2). Fit indices, entropy (.964), and average posterior probabilities of participants being correctly (98–99%) and incorrectly (0–1%) assigned to each of the three classes indicated high classification accuracy, specificity, and sufficient statistical power. Classes reflected three subgroups with varying symptom presentations: 1) high global symptoms, reflective of depression, anxiety, and PTSD (*n*=48, 23.8%); 2) Dysphoric Arousal (*n*=96, 47.5%); and 3) Hyperarousal (*n*=58, 28.7%; Supplemental Table 2). Dysphoric Arousal was characterized by a higher probability of endorsing lack of interest/motivation, diminished positive emotions, negative thought patterns, social withdrawal, fatigue, concentration issues, hypervigilance, trouble relaxing, and sleep problems. Hyperarousal was characterized by elevations in irritability, fatigue, concentration issues, and sleep problems, but lower overall symptoms.

**Figure 2.**
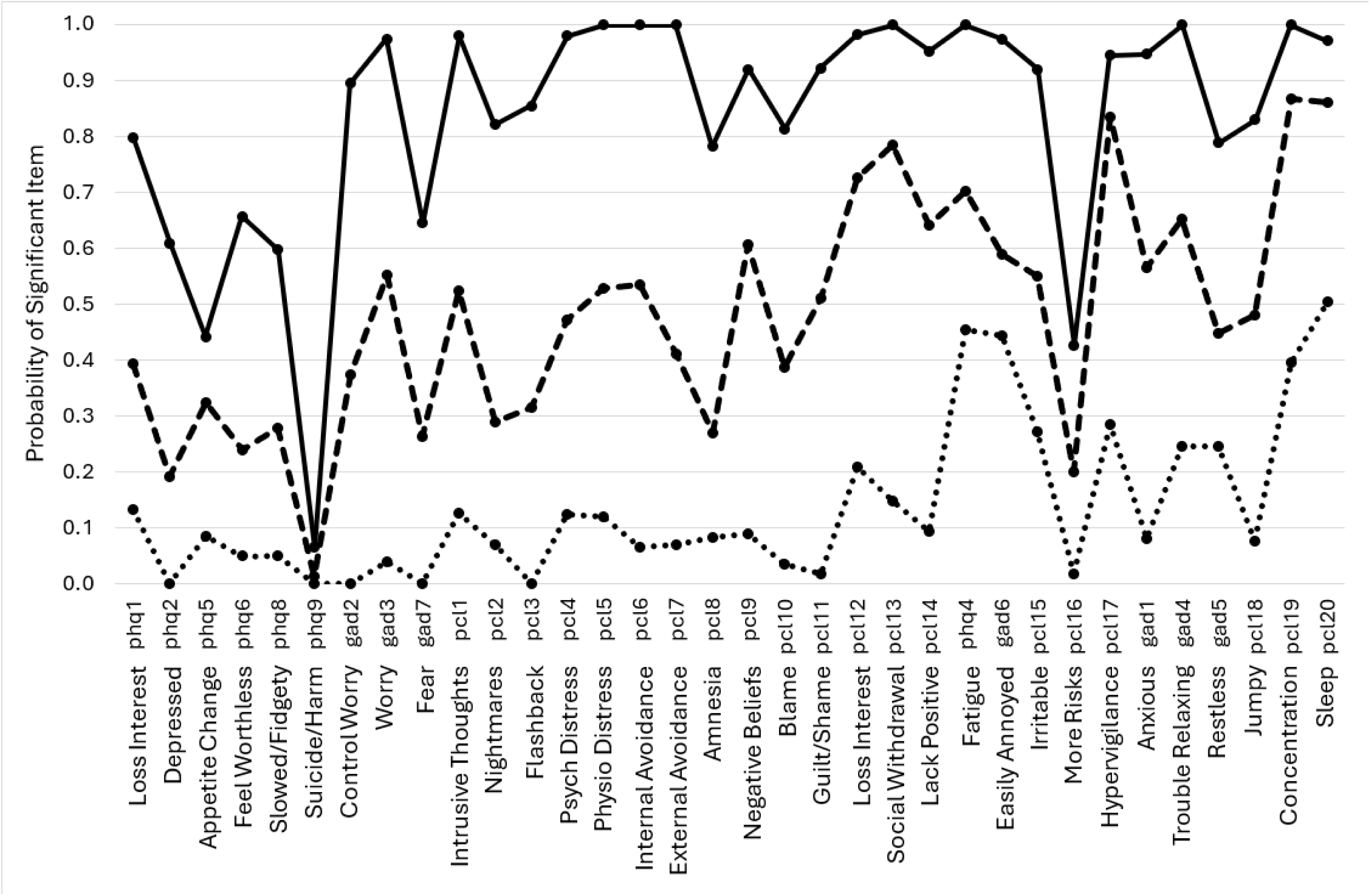
LCA results of three-class solution for OS Mental Health Disruption.

## Discussion

This study provides the first empirical validation of OS as a unitary construct using a representative sample of treatment-seeking active-duty SOF personnel with significant histories of TBI. Nearly nine-out-of-ten (89.1%) participants met criteria for OS with Sleep, Cardiometabolic, Pain, Mental Health, Headaches, and Cognitive Disruptions occurring in more than three-quarters (78%) of participants. Three types of mental health disruptions were identified in OS with Dysphoric Arousal being the predominant presentation. Overall, findings indicate the utility of OS in guiding more targeted, effective, and engaging clinical interventions for the breadth of unique problems common to SOF. Below we summarize findings in more detail and provide actionable recommendations for current and future research, clinical, and policy applications.

### OS Structure

This study aimed to 1) evaluate the empirical validity and structure of OS, 2) identify the core features of OS, and 3) evaluate if OS represents a unique construct that offers further utility beyond existing psychiatric diagnoses. Previous delays in the research development of OS were, in part, due to difficulties attaining a representative sample of operators and methodological difficulties measuring the breadth of multidimensional and multimodal variables relevant to OS. Both issues were addressed here using MOA employed within the PSC and IETP setting (10):

1. Results of both variable-centered (EFA) and person-centered (LCA) statistical approaches identified one factor/class, supporting the validation of OS as a unitary construct.
2. In addition to essential prerequisites of repetitive and chronic exposure to high-tempo tasks that are physically demanding and involve extreme threats to physical safety, exposure to potentially traumatic event(s), and/or TBI, OS was composed of nine core domains: Cardiometabolic, Chronic Localized and Non-Specific Pain, Cognitive, Endocrine, Gastrointestinal, Headaches, Mental Health, Sensory, and Sleep Disruptions (Table 3). Headaches, Mental Health, Pain, Sensory, and Sleep Disruptions accounted for roughly 69% of the total variance in OS outcomes, suggesting ≥ 5 domains may be a suitable criterion for OS. Identifying core features of OS required pruning items that were secondary rather than direct consequences of OS. Substance abuse, marital/family concerns, and intimacy concerns within the original OS framework were considered secondary consequences of existing symptoms like emotion dysregulation, rather than direct consequences of trauma- and TBI-related OS exposures (39). Therefore, they were excluded from the core features of OS to aid the implementation of assessment, treatment, and policy practices.
3. Further analysis indicated that OS is related to other mental health conditions (i.e., anxiety, depression, PTSD), as expected, but was not collinear with these conditions. Accordingly, results suggest that OS reflects a construct related to but distinct from existing psychiatric diagnoses that may offer clinical utility over and above psychiatric diagnoses alone (11). Historically, other diagnoses like PTSD were also criticized as being overly broad (40,41). Nosologists argue that clinical constructs like PTSD and OS, should strive to “carve nature at its joints” (42) describing the core features of the syndrome. Doing so can both help limit comorbidity or overlap with other disorders and help describe what is central to the experience of OS, providing clear and actionable targets for assessment and treatment.

**Table 3.** Changes in revised Operator Syndrome framework.

| Original Operator Syndrome Framework | Revised Operator Syndrome Framework |
| --- | --- |
|  | **Criterion A – exposure prerequisites** |
| Traumatic Brain Injury | Exposure to Trauma and/or Traumatic Brain Injury (TBI) |
|  | Chronic exposure to high-tempo operations |
|  | **Criterion B – need at least 5 out of 9** |
| Chronic Pain, Orthopedic Problems, Headache | Chronic Localized and Non-Specific Pain |
|  | Headaches |
|  | Cardiometabolic Disruption |
| Cognitive Impairments | Cognitive Disruption |
| Endocrine Dysfunction | Endocrine Disruption |
|  | Gastrointestinal Disruption |
| Depression | Mental Health Disruption<br>(e.g., Dysphoric Arousal: fatigue, lack of interest/motivation, numb/blunted positive emotions, social withdrawal, hyperarousal/restless, irritability/anger, sleep disruption) |
| Anxiety |  |
| Anger |  |
| Hypervigilance |  |
| PTSD |  |
| Perceptual System Impairments | Sensory Disruption |
| Sleep Disturbance | Sleep Disruption |
| Substance Abuse | - |
| Marital and Family Concerns | - |
| Intimacy Concerns | - |
|  | **Criterion C - associated features and specifiers** |
| Toxic Exposures and Related Illness | *With Elevated Heavy Metals/Toxins* |
| Military-Civilian Transition | *With Moral Injury, Existential Disruption, and/or Military Transition* |
| Existential Concerns |  |

Preliminary analyses of a smaller subset of participants indicated that Moral Injury (36.2%) and Heavy Metals/Toxins (56.0%) were prevalent in this SOF sample. Although these are preliminary data that require further examination, these domains are relevant to the SOF community and were included in the original OS conceptual framework (5). Therefore, they were retained in the revised OS model. Like posttraumatic stress, moral injury is a heterogeneous construct that presents differently for many individuals (10,29). Moral injury is the result of direct, witnessed, or second-hand knowledge of acts that conflict with one’s deeply held moral beliefs (29). For some, this stems from acts (e.g., murder, assault), but for many, this is the result of perceived injustices or broader inconsistencies between events and one’s morally held beliefs or expectations (e.g., 2021 withdrawal from Afghanistan) (43). Such exposures can exacerbate a jaded change in negative thoughts and existential beliefs about oneself, others, and/or the world in general.

Heavy metals and toxins are also relevant to OS as the effects of elevated exposure to heavy metal toxins can mimic stress- and TBI-related symptoms (e.g., cognition, mood, gastrointestinal issues, pain) compounding these issues and complicating recovery (44).

Although heavy metals are found in many components of diet (e.g., fish), supplements (e.g., plant-based protein powders), and environment (e.g., water, soil), SOF personnel can also have unique occupational exposures to these toxins (e.g., foreign environments, explosives, ammunition, combat simulation houses) (45–47). We must also emphasize that total levels of heavy metals/toxins were measured in this study. Therefore, we cannot delineate organic versus inorganic contributions to heavy metal levels, which are important for discerning toxic effects. These preliminary findings require further investigation.

Notably, original OS domains of Moral Injury, Existential Disruption, and Military Transition and Heavy Metals/Toxins were excluded as core features of OS because individuals can experience OS without these components. Although OS can be exacerbated by these domains, the experience of OS is not dependent on them. This parallels the logic for excluding substance abuse, family/interpersonal problems, and intimacy problems from the revised OS model. These issues are certainly related to OS, but they are secondary to the experience of core OS features identified here and, therefore, excluded.

Lastly, EFA identified Cardiometabolic Disruption as the weakest OS domain, and LCA identified Endocrine Disruption as the weakest OS domain. Because of these discrepant results, the authors faced a decision to either exclude or retain both Cardiometabolic and Endocrine Disruption in the OS model. Given the high prevalence and high LCA probability of Cardiometabolic Disruption, and the significant EFA loading of Endocrine Disruption, both were retained in the final OS model because of their respective statistical contributions as well as potential clinical utility.

### Systemic Illness of OS

OS reflects the systemic interactions of multiple injuries that can produce and exacerbate a constellation of symptoms over time. Systemic processes like these have been noted following trauma, TBI, and chronic states of stress (10,12,48). If unmitigated, these systemic processes can produce more complicated health conditions over time (e.g., cardiovascular disease, metabolic syndrome), prompting the call for early comprehensive and integrative intervention practices, such as those within PSC (12). Cardiometabolic and Gastrointestinal Disruptions were common in this SOF sample and added to OS to help reflect these systemic processes and guide relevant clinical assessments and interventions.

A strong relationship between chronic stress, TBI, and both Cardiometabolic and Gastrointestinal Disruption has been well documented (12,13,49,50). The relationships between internal bodily sensations/organs and psychological/neurological processes is called interoception and has received great interest across numerous biological systems (e.g., cardioception, pulmoception, gastroception, uroception) (51,52). Multiple injuries and chronic stress, reflecting increased allostatic load, can dysregulate multiple biological systems and increase risk for insulin resistance, contributing to elevated heart rate, blood pressure, cholesterol, and blood glucose (12,49,53). When sustained over time, these elevations increase risk for cardiovascular disease, hypertension, hyperlipidemia, Type II diabetes, and, in some cases, mortality (12,54).

Similarly, multiple injuries and chronic elevations in stress can contribute to Gastrointestinal Disruptions, reflected by chronic constipation, diarrhea, changes in bowel movement frequency and consistency, and gastroesophageal reflux (55,56). Altered bowel movements and significant abdominal discomfort are often diagnosed as functional gastrointestinal disorders (FGID) related to interoceptive processes and conditions like gastroesophageal reflux disease (GERD) and irritable bowel syndrome (IBS) (51,52,57). SOF personnel are not only exposed to these gastrointestinal effects through stress and TBI, but also through varied environmental exposures, substances, toxins, diet, and changing dietary habits/schedules inherent to their operations and lifestyle (58). Clinical trials have indicated that a moderate “psychobiotic” diet (i.e., prebiotic, polyphenol, anti-inflammatory) can help reduce perceived stress and chronic pain (56,58,59), suggesting the importance of nutrition and gastrointestinal health as one target in an integrative OS treatment.

### Closer Examination of Mental Health Disruption

To help curb critiques that contend OS is merely reflective of existing psychiatric diagnoses like PTSD, we conducted a detailed examination of Mental Health Disruption, including depressive, anxious, and posttraumatic stress symptoms. The goal of this examination was to determine if OS symptoms were distinct from existing psychiatric diagnoses and to identify precise presentations that may provide specific targets beyond the original OS framework for increasingly effective and engaging interventions.

Three types of presentations were identified: 1) high global symptoms, reflective of traditional presentations of depression, anxiety, and PTSD; 2) Dysphoric Arousal; and 3) Hyperarousal. Most participants (76.2%) presented with Dysphoric Arousal or Hyperarousal, with fewer presenting with traditional forms of depression, anxiety, and PTSD. Dysphoric Arousal was the predominant (47.5%) presentation, characterized by dysphoria (i.e., lack of interest/motivation, diminished positive emotions, negative thought patterns, social withdrawal, fatigue) and arousal (i.e., concentration issues, hypervigilance, trouble relaxing, sleep problems). Hyperarousal is one of four clusters of posttraumatic stress-related symptoms characterized by irritability, fatigue, hypervigilance, concentration issues, and sleep problems (11). For both presentations, intrusive symptoms (e.g., thoughts/memories of the traumatic event[s], flashbacks, reactivity) and avoidance symptoms were less prevalent, helping to differentiate these symptom presentations from PTSD.

Accordingly, Dysphoric Arousal is not well-captured by current psychiatric diagnoses. This presentation is considered subthreshold or unspecified, illustrating an ongoing issue of representation of SOF Mental Health Disruptions for healthcare services and insurance coverage. Despite lack of formal recognition, Dysphoric Arousal is well-represented in the literature (60–62). Moreover, Hyperarousal describes a foundational aspect of OS, in part, because SOF are trained to have heighted situational awareness during training and deployments. However, when overgeneralized to everyday situations, hyperarousal and hypervigilance can become burdensome and impair one’s ability to enjoy the daily activities of civilian life. Overgeneralized hyperarousal can also lead to exacerbations in other Mental Health Disruptions over time, providing a target for intervention (50,60).

Core features of Dysphoric Arousal are emotional blunting and dysphoria with hyperarousal (60–62). Individuals feel on-edge, restless, and/or a sense of urgency, while concurrently feeling stuck, unmotivated, fatigued, and experience diminished interest and pleasure in most things. They also have an increase in negative thought patterns, accompanied by a more global alteration in beliefs about oneself, others, and the world in general, which can also be related to moral injury. Positive emotions are blunted and individuals may struggle to hold onto fleeting positive moments and emotions. Over time, this may cause them to withdraw from situations that no longer bring enjoyment and are no longer positively reinforcing. Adynamia (lack of motivation/drive), resulting from TBI and Endocrine Disruption, compounds Dysphoric Arousal (63). In turn, this can exacerbate issues of fatigue and lack of enjoyment or pleasure, contributing to social withdrawal and distancing, isolation and loneliness, and make engagement in activities more difficult and exhausting. Consequently, these symptoms can contribute to substance use difficulties, sexual dysfunction, and straining family and other interpersonal relationships.

### Recommendations for OS

Current findings support nine core domains of OS. The presence of ≥ 5 domains indicated good sensitivity in detecting OS and specificity in parsing OS apart from related psychiatric diagnoses. Previously, Frueh et al. (5) included TBI in congruence with these domains. However, TBI is a catalyst for these domain-specific problems (the injury) just as trauma is the catalyst for posttraumatic stress symptoms. To accommodate and align with current policy and implementation practices, we have organized OS using existing diagnostic frameworks. Like PTSD and TBI, which have exposure criterion, we have added Criterion A for OS, which is a prerequisite for subsequent Criterion B (OS symptoms) and Criterion C (specifiers; Table 3). Criterion A includes:

a. Repetitive and chronic exposure to high-tempo tasks that are physically demanding and involve extreme threats to physical safety; *and*
b. Exposure to potentially traumatic event(s); *and/or*
c. Exposure to events likely to cause traumatic brain injury (TBI).

“Chronic” is a subjective term but is defined by the American Psychiatric Association (APA) as ≥ 2 years (11). These criteria reflect necessary precipitants to OS and help parse OS apart from other injury and stress reactions (e.g., chronic stress without trauma or TBI). With Criterion B reflecting ≥ 5 OS domains, OS is most plainly described as a response to years of sustained exposure to intense, threatening environments, brain injury and/or trauma exposure, and subsequent allostatic load, reflecting the cumulative effect of ongoing stressors and burnout. Lastly, Criterion C was added, including Moral Injury, Existential Disruption, and/or Military Transition and Heavy Metals/Toxins. This criterion is aligned with the APA’s “specifiers.” These are preliminary domains that may not be central to the experience of OS (i.e., one can experience OS without these domains) but are prevalent in this sample and can exacerbate other symptoms of OS as described above. The addition of these specifiers can also help guide clinical intervention.

Despite the term, OS, these exposures and effects are not specific to SOF operators. Just as the term, Legionnaire’s disease does not solely describe members of the American Legion, OS is named after the operators it originally described but has since been generalized to others.

Professions with similar exposures and demands, depending on their duties and environment (e.g., other USSOCOM personnel, conventional military, first-responders), can see similar effects (64). However, these exposures and injuries are common to SOF given their extreme occupational demands and accompanied lifestyle, prompting a need for further investigation and implementation to treatment, performance optimization, and policy.

A detailed commentary on treatment recommendations is beyond the scope of this study. Rather than recommending one treatment modality or another, here, we will briefly comment on the critical and necessary utility of comprehensive and integrative treatments for OS. Integrated treatments are one of the most critical components of successful treatment practices following TBI and multiple injuries (6). VHA PSC programs like IETP are some of the most well-equipped to treat OS and offer significant advantages over other specialty and private clinics, which may offer more siloed care that may fail to assess critical components of OS and require additional referrals to receive comprehensive care. Further detailed treatment recommendations for OS will be offered in future studies.

### Future Directions

There are innumerable future directions for the study and implementation of OS. First, this revised OS model requires ongoing evaluation. Second, the effects of moral injury and inorganic heavy metals/toxin exposures on OS promptly require further study. Third, additional influences on OS should continue to be examined (e.g., biomarkers, cultural/environmental influences, behavioral/personality influences, related health conditions) to provide further precision and guidance for increasingly targeted and effective treatments. Fourth, future studies should assess additional components of Endocrine Disruption to better capture the breadth of systemic effects of OS.

Lastly, the broader objective of this study was to progress OS so that it can eventually be formally recognized in policy and implementation practices on a large scale. OS may be most appropriately recognized as a formal syndrome, much like Gulf War Syndrome, or diagnostic “z-code,” much like moral injury. Z-codes are delineated by the International Classification of Diseases (ICD) and used by medical professionals to document other conditions that may be a focus of clinical attention and may be critical to inform the patient’s care plan (11). Formal inclusion in this capacity can not only assist with patient satisfaction and equitable representation of an underrepresented problem and population but can assist with attaining appropriate healthcare coverage and accommodations. Outside of direct healthcare, adoption of the OS framework may have profound implications for performance optimization among active-duty SOF and similar populations, including more focused attention to TBI prevention, operational tempos, sleep and circadian disruption, endocrinology, and nutrition, as well as mitigation of heavy metals and brain health hazards to the extent possible.

### Limitations

Despite the strengths of this study, there are some limitations. Namely, data for Gastrointestinal Disruption were collected during clinical intake and dichotomously recorded. Future studies should incorporate more sensitive measures of Gastrointestinal Disruption.

Although more women are in command or support roles for USSOCOM, only a handful have passed the strict SOF selection process for operations (65). The current sample of three women approximates the proportion of men to women in SOF, but findings are not representative of SOF women. Future studies should aim to address the unique health needs that may be pertinent to SOF women. Lastly, only preliminary data were available for moral injury and heavy metals/toxins. Further study is required.

## Conclusion

This study provides the first empirical validation of OS as a unitary construct, evidence of the core features of OS, and demonstrates that OS can provide unique utility beyond current psychiatric diagnoses. Several recommendations and guidelines are provided to help direct current applications and future directions for OS. This study also provides evidence of a predominant OS-related mental health presentation – Dysphoric Arousal – that can bring precision to the triage of clinical care for OS, producing increasingly engaging and efficacious treatments for SOF personnel. Although further study is needed, this study helps progress OS in an effort to represent the unique experiences of SOF personnel and the consequences they have incurred through years of service.

## Data Availability

The dataset generated during the current study is not publicly available as it includes VHA protected health information, but select de-identified data may be available from the corresponding author on reasonable request.

## Acknowledgements

We sincerely recognize and thank the significant contributions of our IETP participants, first and foremost, as well as the teams of military personnel and health care coordinators that facilitate their entry into our program and subsequent follow-up care. We also sincerely acknowledge the invaluable contributions of our IETP team, without whom the foundation of this work could not stand: Qiliang Chen, Debbie Cheng, Lindsay Conner, Katrin Cooper, Cristina Dizon, Jamie Eamiguel, Esther Estey, Kathleen Gorman, Adyam Hagos, Yolly Hardin, Rebecca Lehmann, Jennifer Loughlin, Milica Lubic, Emily McCrone, Elisabeth McKenna, Nadja Mencin, Christina Patron, Chenal Roberts, Tiffany Segura, Chetna Singh, Jeffrey Teraoka, Julia Victor, Nina Wakayama, Tiffanie Sim Wong, and Joseph Yang.

## References

1. United States Special Operations Command. USSOCOM Core Activities [Internet]. 2024 [cited 2025 Jun 9]. Available from: https://www.socom.mil/about/core-activities

2. Johnson M. The growing relevance of special operations forces in U.S. military strategy. Comparative Strategy [Internet]. 2006 Sep 1;25(4):273–96. Available from: 10.1080/01495930601028622

3. Congressional Research Service. U.S. Special Operations Forces (SOF): Background and Considerations for Congress [Internet]. Washington, D.C.; 2025 Mar [cited 2025 Oct 5]. Available from: https://crsreports.congress.gov RS21048

4. USSOCOM. United States Special Operation Command Comprehensive Review [Internet]. Tampa; 2020 Jan [cited 2025 Oct 5]. Available from: https://int.nyt.com/data/documenthelper/6736-special-operations-forces-review/c93cf96d67c341b2d2c3/optimized/full.pdf

5. Frueh BC, Madan A, Fowler JC, Stomberg S, Bradshaw M, Kelly K, et al. “Operator syndrome”: A unique constellation of medical and behavioral health-care needs of military special operation forces. The International Journal of Psychiatry in Medicine [Internet]. 2020 Feb 13;55(4):281–95. Available from: 10.1177/0091217420906659

6. Garcia A, Kretzmer TS, Dams-O’Connor K, Miles SR, Bajor L, Tang X, et al. Health conditions among special operations forces versus conventional military service members: A VA TBI Model Systems study. J Head Trauma Rehabil [Internet]. 2022;37(4). Available from: https://journals.lww.com/headtraumarehab/fulltext/2022/07000/health_conditions_among_special_operations_forces.15.aspx

7. Stannard J, Fortington L. Musculoskeletal injury in military Special Operations Forces: A systematic review. BMJ Mil Health [Internet]. 2021 Aug 1;167(4):255. Available from: http://militaryhealth.bmj.com/content/167/4/255.abstract

8. Eapen BC, Jaramillo CA, Tapia RN, Johnson EJ, Cifu DX. Rehabilitation care of combat related TBI: Veterans Health Administration Polytrauma System of Care. Curr Phys Med Rehabil Rep [Internet]. 2013;1(3):151–8. Available from: 10.1007/s40141-013-0023-0

9. French MA, Roemmich RT, Daley K, Beier M, Penttinen S, Raghavan P, et al. Precision rehabilitation: Optimizing function, adding value to health care. Arch Phys Med Rehabil [Internet]. 2022;103(6):1233–9. Available from: https://www.sciencedirect.com/science/article/pii/S0003999322002131

10. Adams SW, Layne CM, Contractor AA, Allwood MA, Armour C, Inslicht SS, et al. The Middle-Out Approach to reconceptualizing, assessing, and analyzing traumatic stress reactions. J Trauma Stress [Internet]. 2023 Dec 4;n/a(n/a). Available from: 10.1002/jts.23005

11. American Psychiatric Association. Diagnostic and statistical manual of mental disorders. 5th-TR ed. Author; 2022.

12. Adams SW, Allwood MA. Parallel processes of posttraumatic stress and cardiometabolic dysfunction: A systemic illness of traumatic stress. Health Psychol [Internet]. 2024;43(5):365–75. Available from: http://europepmc.org/abstract/MED/38127510

13. Koenen KC, Sumner JA, Gilsanz P, Glymour MM, Ratanatharathorn A, Rimm EB, et al. Post-traumatic stress disorder and cardiometabolic disease: Improving causal inference to inform practice. Psychol Med [Internet]. 2016/10/04. 2017 Jan;47(2):209–25. Available from: https://pubmed.ncbi.nlm.nih.gov/27697083

14. Grundy S, Cleeman J, Daniels S, Donato K, Eckel R, Franklin B, et al. Diagnosis and management of the metabolic syndrome: an American Heart Association/ National Heart, Lung, and Blood Institute scientific statement: executive summary. Crit Pathw Cardiol. 2005;4:198–203.

15. Graham K, Searle A, Van Hooff M, Lawrence-Wood E, McFarlane A. The value of physical symptoms in screening for posttraumatic stress disorder in the military. Assessment [Internet]. 2019 Jul 21;27(6):1139–50. Available from: 10.1177/1073191119864662

16. Osman A, Barrios FX, Kopper BA, Hauptmann W, Jones J, O’Neill E. Factor structure, reliability, and validity of the pain catastrophizing scale. J Behav Med [Internet]. 1997;20(6):589–605. Available from: 10.1023/A:1025570508954

17. Amtmann D, Cook KF, Jensen MP, Chen WH, Choi S, Revicki D, et al. Development of a PROMIS item bank to measure pain interference. Pain [Internet]. 2010;150(1):173–82. Available from: https://www.sciencedirect.com/science/article/pii/S0304395910002630

18. Cella D, Lai JS, Nowinski CJ, Victorson D, Peterman A, Miller D, et al. Neuro-QOL: Brief measures of health-related quality of life for clinical research in neurology. Neurology [Internet]. 2012;78(23):1860–7. Available from: http://europepmc.org/abstract/MED/22573626

19. Helm-Estabrooks N. Cognitive Linguistic Quick Test. San Antonio, TX: PsychCorp; 2001.

20. Ramsay MC, Reynolds CR. Separate digits tests: A brief history, a literature review, and a reexamination of the factor structure of the test of memory and learning (TOMAL). Neuropsychol Rev [Internet]. 1995;5(3):151–71. Available from: 10.1007/BF02214760

21. Barnett N, Ljubic M, Chung J, Capizzi A. Testosterone and neurobehavioral outcomes in special operations forces military with multiple mild traumatic brain injury. NeuroRehabilitation [Internet]. 2024 Jul 9;55(3):271–9. Available from: https://journals.sagepub.com/action/showAbstract

22. Lunenfeld B, Nieschlag E, Lunenfeld B, Nieschlag E. Testosterone therapy in the aging male. The Aging Male [Internet]. 2007 Jan 1;10(3):139–53. Available from: 10.1080/13685530701485998

23. Rendas-Baum R, Yang M, Varon SF, Bloudek LM, DeGryse RE, Kosinski M. Validation of the Headache Impact Test (HIT-6) in patients with chronic migraine. Health Qual Life Outcomes [Internet]. 2014;12(1):117. Available from: 10.1186/s12955-014-0117-0

24. Stewart WF, Lipton RB, Dowson AJ, Sawyer J. Development and testing of the Migraine Disability Assessment (MIDAS) Questionnaire to assess headache-related disability. Neurology [Internet]. 2001 Mar 1;56(suppl_1):S20–8. Available from: 10.1212/WNL.56.suppl_1.S20

25. Keil DE, Berger-Ritchie J, McMillin GA. Testing for toxic elements: A focus on arsenic, cadmium, lead, and mercury. Lab Med [Internet]. 2011 Dec 1;42(12):735–42. Available from: 10.1309/LMYKGU05BEPE7IAW

26. Rutter LA, Brown TA. Psychometric properties of the Generalized Anxiety Disorder Scale-7 (GAD-7) in outpatients with anxiety and mood disorders. J Psychopathol Behav Assess [Internet]. 2017;39(1):140–6. Available from: 10.1007/s10862-016-9571-9

27. Manea L, Gilbody S, McMillan D. Optimal cut-off score for diagnosing depression with the Patient Health Questionnaire (PHQ-9): A meta-analysis. Can Med Assoc J [Internet]. 2012 Feb 21;184(3):E191. Available from: http://www.cmaj.ca/content/184/3/E191.abstract

28. Bovin MJ, Marx BP, Weathers FW, Gallagher MW, Rodriguez P, Schnurr PP, et al. Psychometric properties of the PTSD Checklist for Diagnostic and Statistical Manual of Mental Disorders-Fifth Edition (PCL-5) in veterans. Psychol Assess [Internet]. 2016;28(11):1379–91. Available from: http://europepmc.org/abstract/MED/26653052

29. Maguen S, Griffin BJ, Pietrzak RH, McLean CP, Hamblen JL, Norman SB. Using the Moral Injury and Distress Scale to identify clinically meaningful moral injury. J Trauma Stress [Internet]. 2024 Aug 1;37(4):685–96. Available from: 10.1002/jts.23050

30. Obuchi C, Kaga K. Development of a questionnaire to assess listening difficulties in adults with auditory processing disorder. Hearing Balance Commun [Internet]. 2020;18(1). Available from: https://journals.lww.com/hbcm/fulltext/2020/18010/development_of_a_questionnaire_to_assess_listening.5.aspx

31. Power Laura, Murray Katherine, Szmulewicz David J. Characteristics of assessment and treatment in Benign Paroxysmal Positional Vertigo (BPPV). Journal of Vestibular Research [Internet]. 2019 Dec 13;30(1):55–62. Available from: https://journals.sagepub.com/action/showAbstract

32. Tamber AL, Wilhelmsen KT, Strand LI. Measurement properties of the Dizziness Handicap Inventory by cross-sectional and longitudinal designs. Health Qual Life Outcomes [Internet]. 2009;7(1):101. Available from: 10.1186/1477-7525-7-101

33. Curran K, Holland G, Redfern MS, Cham R. Comparing sensory organization test measures of the Bertec® Balance Advantage® CDP/IVRTM and NeuroCom® Smart EquiTest® computerized dynamic posturography systems in young and older healthy adults. Gait Posture [Internet]. 2025;121:308–14. Available from: https://www.sciencedirect.com/science/article/pii/S0966636225002309

34. Morin CM, Belleville G, Bélanger L, Ivers H. The Insomnia Severity Index: Psychometric indicators to detect insomnia cases and evaluate treatment response. Sleep [Internet]. 2011 May 1;34(5):601–8. Available from: 10.1093/sleep/34.5.601

35. Nylund-Gibson K, Choi AY. Ten frequently asked questions about latent class analysis. Transl Issues Psychol Sci. 2018;4(4):440–61.

36. Nylund KL, Asparouhov T, Muthén BO. Deciding on the number of classes in latent class analysis and growth mixture modeling: A monte carlo simulation study. Struct Equ Modeling [Internet]. 2007 Oct 23;14(4):535–69. Available from: 10.1080/10705510701575396

37. Visser M, Depaoli S. A guide to detecting and modeling local dependence in latent class analysis models. Struct Equ Modeling [Internet]. 2022 Nov 2;29(6):971–82. Available from: 10.1080/10705511.2022.2033622

38. Williams B, Onsman Andrys, Brown Ted. Exploratory factor analysis: A five-step guide for novices. Australasian Journal of Paramedicine [Internet]. 2010 Jan 1;8:1–13. Available from: 10.33151/ajp.8.3.93

39. Smith LL, Yan F, Charles M, Mohiuddin K, Tyus D, Adekeye O, et al. Exploring the link between substance use and mental health status: What can we learn from the self-medication theory? J Health Care Poor Underserved [Internet]. 2017;28(2S):113–31. Available from: http://europepmc.org/abstract/MED/28458268

40. Spitzer RL, First MB, Wakefield JC. Saving PTSD from itself in DSM-V. J Anxiety Disord. 2007;21(2):233–41.

41. McNally RJ. Can we fix PTSD in DSM-V? Depress Anxiety. 2009;26(7):597–600.

42. Zachar P, Kendler KS. The philosophy of nosology. Annu Rev Clin Psychol [Internet]. 2017 May 8;13(1):49–71. Available from: 10.1146/annurev-clinpsy-032816-045020

43. Bruce MJ, Little DM, Hernandez-Tejada MA, Acierno R. Posttraumatic stress disorder treatment outcomes for events related to institutional betrayal. J Trauma Stress [Internet]. 2025 Jul 6;n/a(n/a). Available from: 10.1002/jts.23187

44. Hoisington AJ, Stearns-Yoder KA, Kovacs EJ, Postolache TT, Brenner LA. Airborne exposure to pollutants and mental health: A review with implications for United States veterans. Curr Environ Health Rep [Internet]. 2024;11(2):168–83. Available from: 10.1007/s40572-024-00437-8

45. Consumer Reports. Heavy Metals in Protein Supplements. Consum Rep [Internet]. 2025 Oct 14 [cited 2025 Oct 15];1–4. Available from: https://article.images.consumerreports.org/image/upload/v1760108748/prod/content/dam/CRO-Images-2025/Special%20Projects/Consumer-Reports-Protein-Powders-and-Shakes-Contain-High-Levels-of-Lead-Methodology-Test-Results.pdf

46. Vandebroek E, Haufroid V, Smolders E, Hons L, Nemery B. Occupational Exposure to Metals in Shooting Ranges: A Biomonitoring Study. Saf Health Work [Internet]. 2019;10(1):87–94. Available from: https://www.sciencedirect.com/science/article/pii/S2093791118300192

47. Marshall TM, Colvin KL, Nevin R, Macrellis J, Dardia GP. Neurotoxicity Associated with Traumatic Brain Injury, Blast, Chemical, Heavy Metal and Quinoline Drug Exposure. Alternative Therapies in Health & Medicine [Internet]. 2019 Jan 1;25(1):28–34. Available from: https://research.ebsco.com/linkprocessor/plink?id=6fbda224-ed2c-3247-8f95-e77e8d684cf0

48. McFarlane AC. Post-traumatic stress disorder is a systemic illness, not a mental disorder: Is Cartesian dualism dead? Medical Journal of Australia. 2017;206(6):248–9.

49. Guidi J, Lucente M, Sonino N, Fava GA. Allostatic load and its impact on health: A systematic review. Psychother Psychosom [Internet]. 2020 Dec 28;90(1):11–27. Available from: 10.1159/000510696

50. Edmondson D, von Känel R. Post-traumatic stress disorder and cardiovascular disease. Lancet Psychiatry. 2017;4(4):320–9.

51. Lovelace JW, Ma J, Augustine V. Defining cardioception: Heart-brain crosstalk. Neuron [Internet]. 2024 Nov 20;112(22):3671–4. Available from: 10.1016/j.neuron.2024.10.009

52. Verdonk C, Ajijola OA, Khalsa SS. Toward a multidisciplinary neurobiology of interoception and mental health. Curr Opin Neurobiol [Internet]. 2025;94:103084. Available from: https://www.sciencedirect.com/science/article/pii/S0959438825001151

53. Janssen J. New Insights into the Role of Insulin and Hypothalamic-Pituitary-Adrenal (HPA) Axis in the Metabolic Syndrome. Int J Mol Sci [Internet]. 2022 [cited 2025 Oct 7];23(15):8178. Available from: 10.3390/ijms23158178

54. Parker HW, Abreu AM, Sullivan MC, Vadiveloo MK. Allostatic load and mortality: A systematic seview and meta-analysis. Am J Prev Med [Internet]. 2022;63(1):131–40. Available from: https://www.sciencedirect.com/science/article/pii/S0749379722001167

55. Ma L, Yan Y, Webb RJ, Li Y, Mehrabani S, Xin B, et al. Psychological stress and gut microbiota composition: A systematic review of human studies. Neuropsychobiology [Internet]. 2023 Sep 6;82(5):247–62. Available from: 10.1159/000533131

56. Berding K, Bastiaanssen TFS, Moloney GM, Boscaini S, Strain CR, Anesi A, et al. Feed your microbes to deal with stress: A psychobiotic diet impacts microbial stability and perceived stress in a healthy adult population. Mol Psychiatry [Internet]. 2023;28(2):601–10. Available from: 10.1038/s41380-022-01817-y

57. Drossman DA. Functional gastrointestinal disorders: History, pathophysiology, clinical features, and Rome IV. Gastroenterology [Internet]. 2016 May 1;150(6):1262–1279.e2. Available from: 10.1053/j.gastro.2016.02.032

58. Sayers B, Wijeyesekera A, Gibson G. Exploring the potential of prebiotic and polyphenol-based dietary interventions for the alleviation of cognitive and gastrointestinal perturbations associated with military specific stressors. J Funct Foods [Internet]. 2021;87:104753. Available from: https://www.sciencedirect.com/science/article/pii/S1756464621004023

59. Costa A, Lucarini E. Treating chronic stress and chronic pain by manipulating gut microbiota with diet: Can we kill two birds with one stone? Nutr Neurosci [Internet]. 2025 Feb 1;28(2):221–44. Available from: 10.1080/1028415X.2024.2365021

60. Adams SW, Possemato K, Kuhn E. Identifying transdiagnostic and multidimensional prognostic indicators among veterans with PTSD symptoms in brief integrated care settings. Psychol Trauma. 2024;14:1–9.

61. Elhai JD, Biehn TL, Armour C, Klopper JJ, Frueh BC, Palmieri PA. Evidence for a unique PTSD construct represented by PTSD’s D1–D3 symptoms. J Anxiety Disord [Internet]. 2011;25(3):340–5. Available from: http://www.sciencedirect.com/science/article/pii/S0887618510002124

62. Pugach CP, Adams SW, Wisco BE, Pietrzak RH. Identifying transdiagnostic traumatic stress reactions in U.S. military veterans: A nationally representative study. J Trauma Stress [Internet]. 2025 Apr 1;38(2):259–71. Available from: 10.1002/jts.23119

63. Palmisano S, Fasotti L, Bertens D. Neurobehavioral Initiation and Motivation Problems After Acquired Brain Injury. Front Neurol [Internet]. 2020;Volume 11-2020. Available from: https://www.frontiersin.org/journals/neurology/articles/10.3389/fneur.2020.00023

64. Frueh BC. Firefighter Syndrome: Addressing long-term psychological, physical risks. University of Hawai’i News [Internet]. 2023 Aug 29 [cited 2025 Oct 6]; Available from: https://www.hawaii.edu/news/2023/08/29/firefighter-syndrome-long-term-psychological-physical-risks/

65. Defense Advisory Committee on Women in the Services. Defense Advisory Committee on Women in the Services 2024 Annual Report [Internet]. Washington, D.C.; 2024 Dec [cited 2025 Oct 6]. Available from: https://dacowits.defense.gov/Portals/48/Documents/DACOWITS%20Annual%20Report%202024%20Web%20Version.pdf

